# Mapping patient interactions in psychiatric presentations to a tertiary emergency department

**DOI:** 10.1101/2023.05.17.23290083

**Authors:** Michael H McCullough, Michael Small, Binu Jayawardena, Sean Hood

**Affiliations:** School of Computing, The Australian National University, Acton, ACT, Australia; Eccles Institute of Neuroscience, John Curtin School of Medical Research, The Australian National University, Acton, ACT, Australia; School of Physics, Mathematics & Computing, The University of Western Australia, Crawley, WA, Australia; Mineral Resources, Commonwealth Scientific and Industrial Research Organisation, Kensington, WA, Australia; North Metropolitan Health Service, Government of Western Australia, WA, Australia; Division of Psychiatry, UWA Medical School, The University of Western Australia, Crawley, WA, Australia

## Abstract

Reliable assessment of suicide and self-harm risk in emergency medicine is critical for effective intervention and treatment of patients affected by mental health disorders. Teams of clinicians are faced with the challenge of rapidly integrating medical history, wide-ranging psychosocial factors, and real-time patient observations to inform diagnosis, treatment and referral decisions. Patient outcomes therefore depend on the reliable flow of information though networks of clinical staff and information systems. We studied information flow at a systems-level in a tertiary hospital emergency department using network models and machine learning. Data were gathered by mapping trajectories and recording clinical interactions for patients at suspected risk of suicide or self-harm. A network model constructed from the data revealed communities closely aligned with underlying clinical team structure. By analysing connectivity patterns in the network model we identified a vulnerability in the system with the potential to adversely impact information flow. We then developed an algorithmic strategy to mitigate this risk by targeted strengthening of links between clinical teams. Finally, we investigated a novel application of machine learning for distinguishing specific interactions along a patient’s trajectory which were most likely to precipitate a psychiatric referral. Together, our results demonstrate a new framework for assessing and reinforcing important information pathways that guide clinical decision processes and provide complimentary insights for improving clinical practice and operational models in emergency medicine for patients at risk of suicide or self-harm.

## INTRODUCTION

Suicide is a major global public health issue causing over 700 000 deaths per year, often with far-reaching impacts on families and communities that can persist well-beyond each individual tragedy [1, 2]. In addition, the prevalence of suicide and suicidal ideation creates considerable economic burden for society (estimated at over US$90 billion in the USA alone in 2013 [3]) and has been linked to increasing healthcare costs [2].

It has been estimated that as many as 77% of individuals who die by suicide will have made contact with a primary care provider in the year prior to their death [4, 5], and up to 10-20% will have visited an emergency department (ED) within 1-2 months prior [6, 7, 2]. EDs are an important and often primary point of access for mental health support services [8, 9, 10] and therefore provide and opportunity for suicide-risk screening and prevention [2]. However, the population of individuals affected by suicidal behaviours is highly heterogeneous and poses significant challenges for risk assessment and clinical management, especially in emergency settings [11].

Mental health crisis presentations account for 4-10% of ED presentations [12] and are growing in number [13, 14, 15, 16]. This increases strain on EDs [13] and impacts patient flow because mental health presentations typically take longer to assess and staff often report feeling ill-equipped to deal with these patients [12]. Further, EDs are widely understood to be challenging environments for patients affected by mental health issues for reasons including long wait times, noise, lack of privacy, harsh lighting, and negative attitudes of staff [17, 18]. These and other factors result in a predominantly negative experience of acute care settings for mental health patients [19]. This is particularly problematic for patients affected by suicidal behaviours because negative experiences of treatment may increase self-harm risk [20].

While clinical management of suicide-risk patients in emergency settings has been prioritised in many national and international suicide prevention strategies [20] current research into urgent emergency care models for mental health patients is limited [12]. Notably, patient journeys through EDs for mental health presentations are not well understood [21], and there are only a few studies attempting to describe the care pathway and interactions with medical professionals in detail [20]. Multiple studies also report considerable inconsistencies and discrepancies in the clinical practice guidelines and service delivery models for emergency mental health care in the USA, UK, Australia and New Zealand [22, 23, 11, 24, 25, 26, 27]. As a result, there have been calls for further research into methods for monitoring and evaluating the implementation of clinical practice guidelines to improve patient experience and treatment outcomes [11, 20, 28].

We addressed these challenges in the present study by developing a novel quantitative framework for evaluating patient journeys through the ED via statistical and algorithmic approaches from the fields of network science and machine learning. Using observational data collected within a tertiary hospital ED we constructed a data-driven network model of interactions between patients, associates, emergency services, clinical staff and information systems for presentations with suspected risk of suicide or self-harm. We analysed the network model to investigate the flow of clinical information at a systems-level and identified properties of the operational structure in the ED that might adversely impact patient care. Further, we explored patient pathways as dynamic networks to understand processes of clinical decision making and referrals as occurring in practice. Together, this work demonstrates the capability of our new quantitative framework for evaluating models of mental health care in emergency medicine. The additional insights afforded by our approach have the potential to guide improvements in clinical practice and operational models to enhance treatment outcomes for patients with suicide or self-harm risk.

## RESULTS

### Characterising patient trajectories as sequences of clinical interactions

We observed patient trajectories and recorded clinical interactions in a tertiary hospital ED for patients presenting with suspected suicidal ideation or self-harm risk (n=43 patients; n=272 interactions; see Methods). To map possible trajectories from the point of presentation to referral or discharge we constructed a patient trajectory network using only the observational data (Figure 1A). This network captured most of the transitions expected based on the operational structure of the ED. This suggests that our data comprised a representative collection of possible patient trajectories. Discharge against medical advice was observed only once. This occurred while the patient was in the care of the Emergency Medical team, but such events are also possible at other points along a patient trajectory. The most common mode of presentation was by ambulance, police or a combination of these (Figure 1B). The median trajectory time per patient was 1.5 hours, 95% CI [0.3, 3.8] (Figure 1C; trajectory time is defined as the duration between the first and last observation made for each patient). Along each trajectory we captured a median of 5 interactions, 95% CI [2, 12] (Figure 1D), involving a median of 4 different types of clinical staff, 95% CI [2, 5] (Figure 1E).

**Figure 1:**
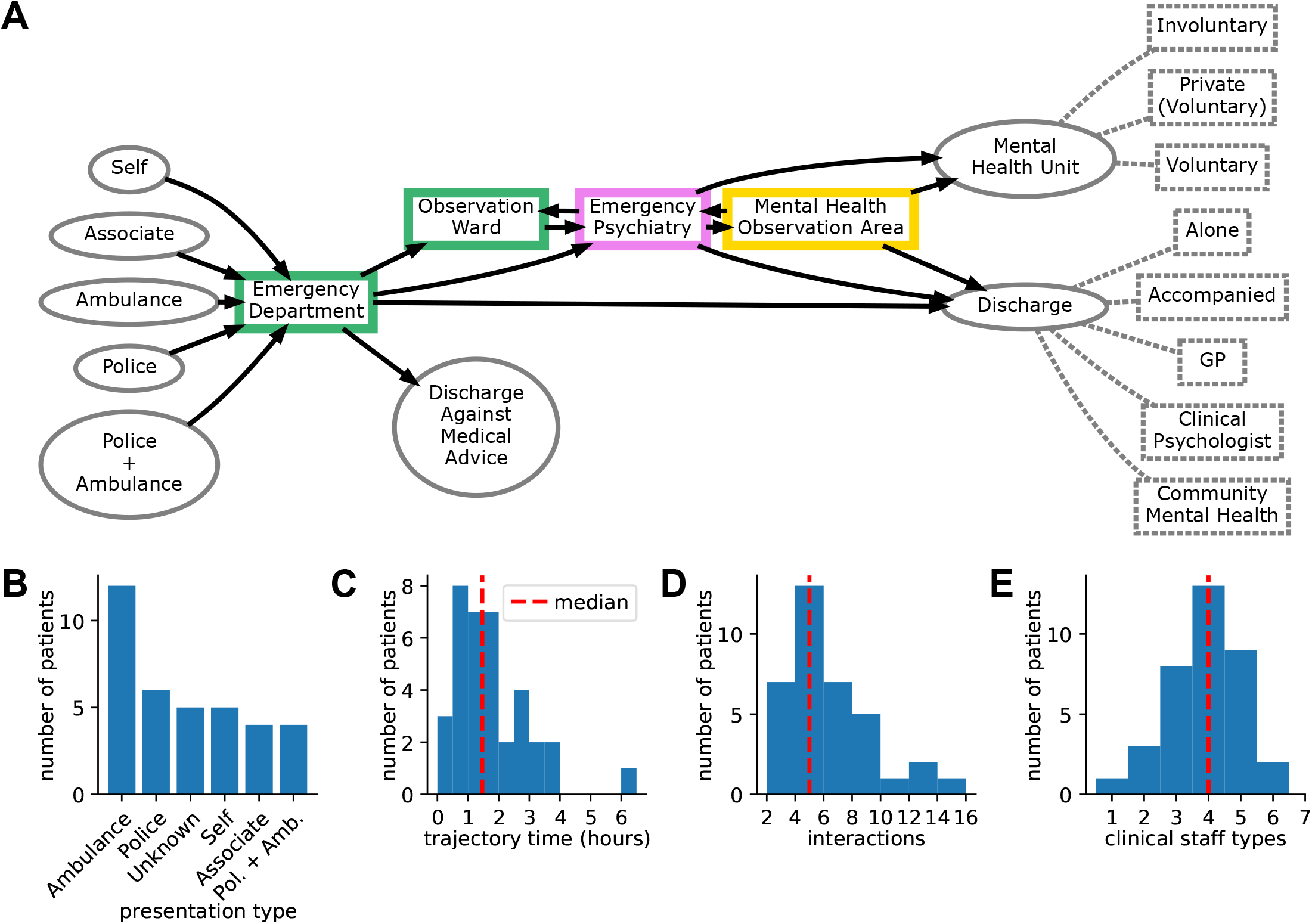
Mapping trajectories through an ED for patients at risk of suicide or self-harm: (A) A network representation of possible patient trajectories constructed from observations from a total of n=43 patients (see Methods). There were n=36 observed within in the ED (including the Observation Ward and Emergency Psychiatry), and n=7 observed only within the Mental Health Observation Area (MHOA). Solid arrows show the observed directional transitions from presentation through the different clinical teams to the point of discharge. The nature of discharge or referral is indicated by the annotations with dashed grey lines. Histograms of (B) presentation type, (C) trajectory time which was the total time between the first and last observation in each patient trajectory, (D) the number of interactions observed per patient, (E) the number of types of clinical staff involved per patient.

### A network model of interactions reflects clinical team structure and reveals agents important for information flow

To investigate the flow of clinical information in the ED we constructed a network model of the of the interactions between agents (i.e., the patient, doctors, or nurses etc.) and clinical information systems as observed along the combined set of patient trajectories in the data (Figure 2A; see Methods). Minimal assumptions were imposed only to ensure that forbidden interactions were not erroneously included in the network (e.g., a hard-copy patient file cannot directly interact with a digital records database). The network edges can be interpreted as communication channels for clinical information that would inform patient diagnosis and treatment. Applying a community detection algorithm [29] to the network reveals a division between the Emergency Medical team and Emergency Psychiatry team based only on the patterns of communications and interactions in the data. The respective internal reporting structures of these teams presumably contributes to this division. However, patient outcomes are likely dependent on effective and reliable communication between teams.

**Figure 2:**
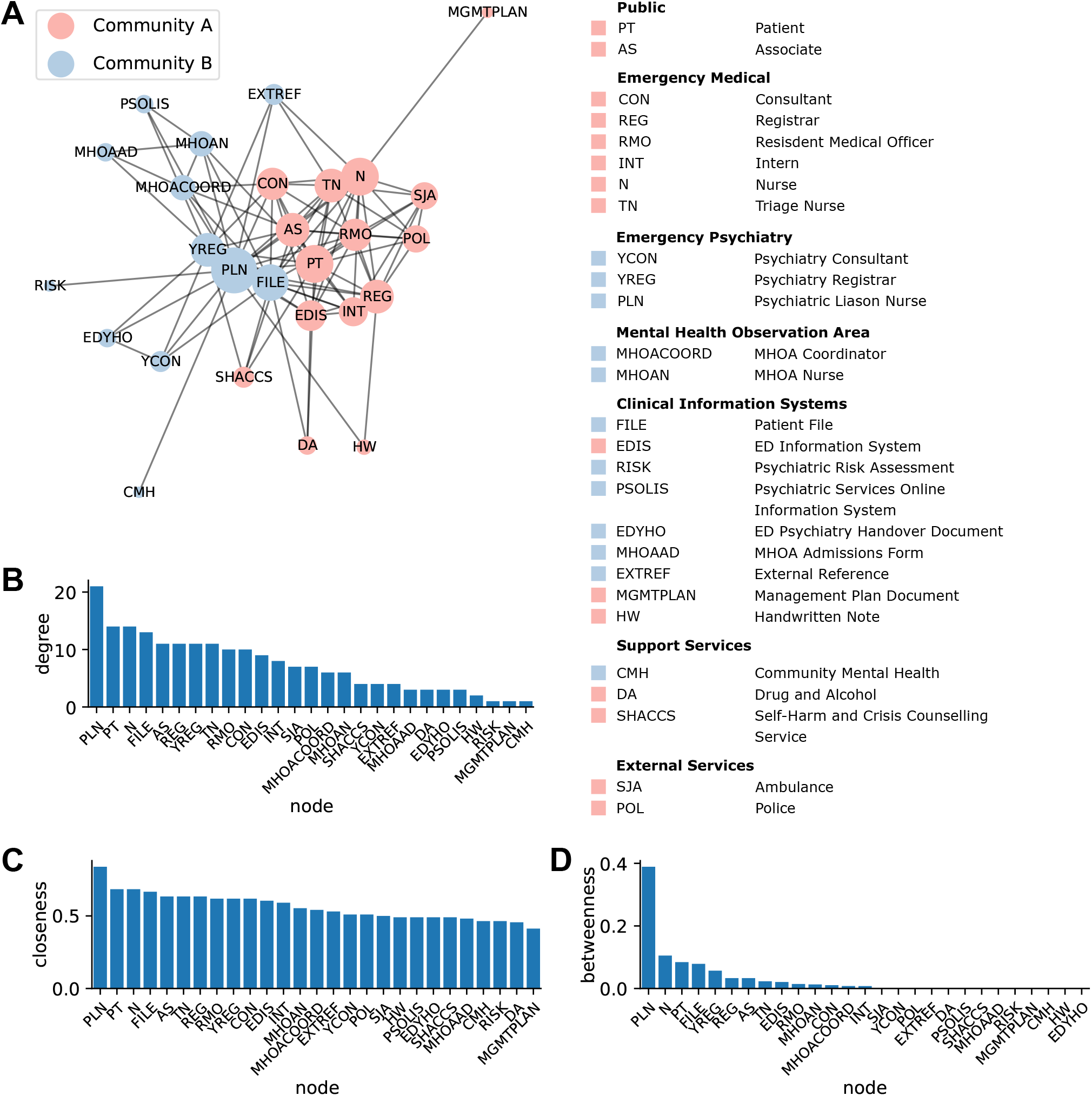
The interaction network comprises two communities divided between the Emergency Medical and Emergency Psychiatry teams and their respective clinical information systems: (A) The interaction network of clinical staff, support services, external services and information systems constructed from observations of n=213 interactions for patient trajectories in the ED (see Methods). Each node represents one of the listed agents. Node size reflects the relative number of instances with which each agent was observed where larger nodes were observed more often. Edges are unweighted and undirected, and represent the observation of at least one interaction between a pair of agents in the data. Two communities were identified by greedy modularity maximisation. Community membership is indicated on both the network visualisation and the glossary of agents. The network layout was generated using a force-directed graph algorithm. (B) Degree centrality, (C) closeness centrality and (D) betweenness centrality of all nodes in the network show high importance of the Psychiatric Liason Nurse (PLN).

Therefore, we next computed measures of node centrality [30] to quantify the importance of individual agents in the interaction network with respect to information flow. Node degree quantifies the direct connectedness and activeness of a node. The psychiatric liaison nurse (PLN), patient, nurse, and patient file were most central in the interaction network by node degree (Figure 2B). This result is unsurprising for the patient, patient file and the nurse given that the patient is the focus of the interactions, the patient file is the primary clinical record, and nurses perform regular observations of the patient. However, it is not immediately apparent as to why the PLN had the highest node degree. Closeness centrality quantifies how central a given node is within the overall structure of the network. The PLN also had the highest closeness centrality (Figure 2C). Values of this measure for all other nodes were relatively consistent. Betweenness centrality quantifies how essential a given node is for transport of information across the network. By this metric the PLN again had the highest centrality, with a value more than four times greater than the agent with the next highest betweenness (Figure 2D). Overall, these results suggest that the PLN is a highly active and connected agent in the ED, and may play a crucial role in communicating clinical information between other agents and treating teams.

### Assessing network vulnerabilities and reducing potential impacts via targeted algorithmic addition of edges

The high node degree and betweenness centrality of the PLN indicates a potential network vulnerability. If the function of the PLN was compromised this might adversely impact the communication of important information between clinical staff. We first sought to establish whether the high betweenness of the PLN resulted from the specific configuration of the interaction network. The alternative hypothesis was that comparable values would arise by chance in similar networks that were configured randomly. To investigate we generated 1000 randomly shuffled versions of the interaction network using a connected double-edge swap algorithm [31] (Fig 3A; see Methods). Betweenness centrality for the PLN was significantly higher in the true network than for random shuffles (>95^th^ percentile). This was not the case for any other of the ten most central agents by betweenness which implies that the specific configuration of the interaction network may impose a unexpectedly high load on the PLN with respect to information transfer. Furthermore, the shuffling algorithm explicitly preserves the degree of each node when shuffling. Therefore, this result also rules out the possibility that the PLN had unexpectedly high betweenness only because it was highly connected, indicating that the position of the agent in the network is important as well as connectivity.

**Figure 3:**
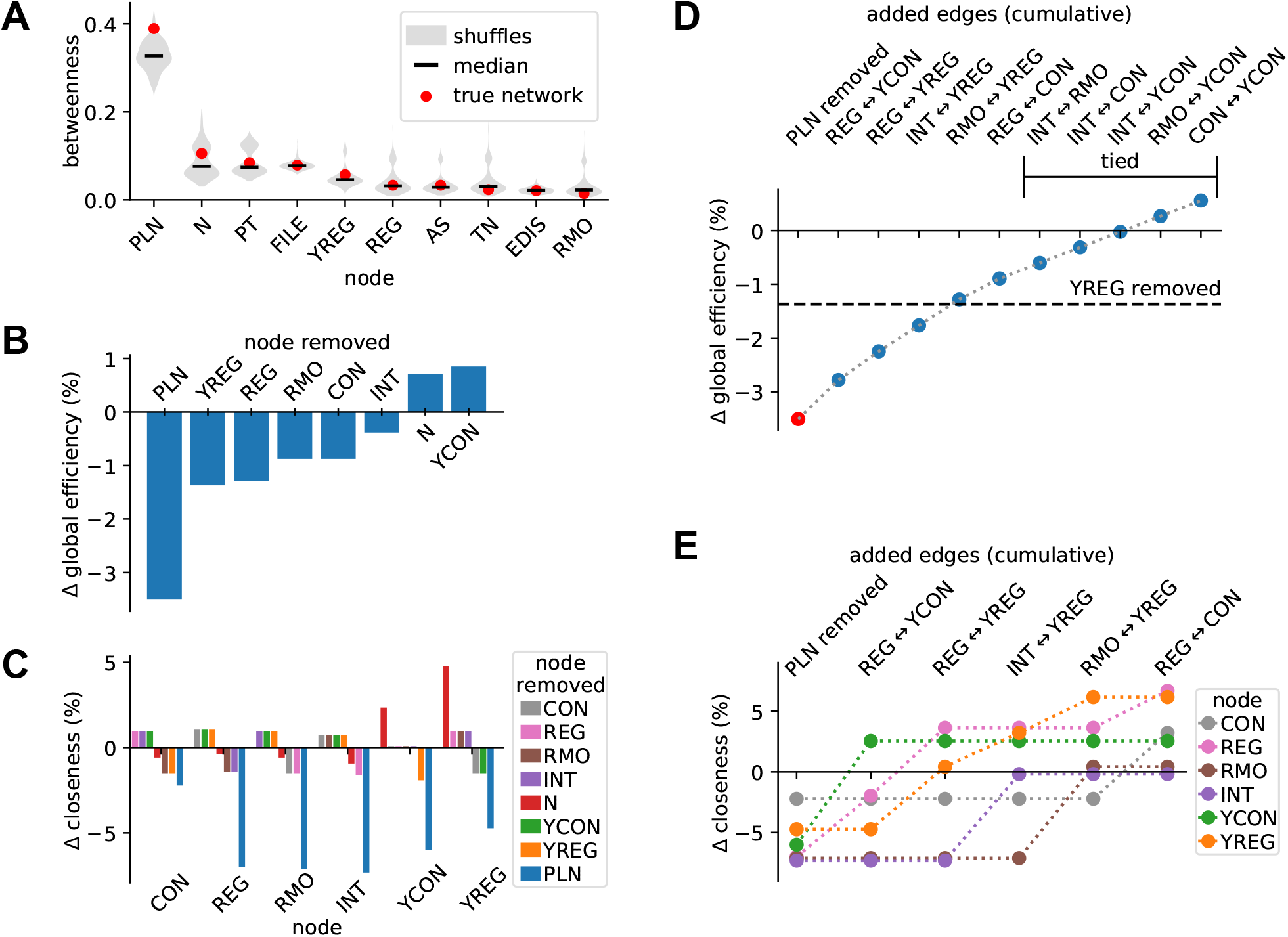
Network vulnerability to compromised function of the PLN is reduced by strengthening links between Emergency Medical and Emergency Psychiatry doctors: (A) The distributions of betweenness centrality shown as violin plots for 1000 random shuffles of the interaction network for the ten nodes with the highest values for this statistic (see Methods). The betweenness for the PLN in the true network is greater than 95^th^ percentile of the shuffles, indicating that this network may be more vulnerable to compromised function of the PLN than expected by chance in similar networks. (B) The change in global efficiency when key clinical staff are removed from the network. (C) The change in closeness centrality for doctors when key clinical staff are removed from the network. (D) Greedy cumulative addition of edges after the removal of the PLN to shows that global efficiency can be restored to a level comparable to the removal of other key clinical staff (dashed line) by the addition of four edges between doctors from the Emergency Medical and Emergency Psychiatry teams. Edges marked as tied contribute equally to the increase in global efficiency regardless of order. (E) Closeness centrality for doctors in the network can be restored by the same greedy addition of edges.

Next, we investigated how information flow might be impacted if the function of the PLN was compromised. Global efficiency measures how efficiently information flows between all pairs of nodes averaged over the network. When the PLN was removed from the network model, global efficiency dropped by more than 3.5% (Figure 3B). This was more than 2.5 times the impact of removing the agent with the next largest impact. The closeness centrality of doctors was reduced considerably more by the removal of the PLN from the network compared with the removal of other clinical staff (Figure 3C). Together, these results indicate that the structure of clinical interactions in the ED may make the system especially vulnerable to compromised function of the PLN. This vulnerability presents a risk to flow of clinical information between agents responsible for decision making along patient trajectories.

We sought a strategy to mitigate this risk by targeted addition of edges between doctors in the Emergency Medical and Emergency Psychiatry teams. We began by removing the PLN from the network then used a greedy algorithm to add edges one at a time to maximise the increase in global efficiency (see Methods). The four edges which contributed most to restoring global efficiency were edges that linked a doctor from the the Emergency Medical team to one from the Emergency Psychiatry team (Fig 3D). The addition of these four edges restored global efficiency to a level comparable to the loss of efficiency which would arise from the compromised function of other clinical staff (Figure 3B). Furthermore, closeness centrality for doctors was fully restored with the exception of the Intern (INT) who required one more edge (Figure 3E). In summary, these results imply that the network vulnerability caused by the high centrality of the PLN could be reduced by increasing communication between doctors from the Emergency Medical and Emergency Psychiatry teams.

### Identifying types of interactions that precipitate clinical handovers with machine learning on interaction networks

We then studied how patterns of interactions influence decision points in patient trajectories. Specifically, we used machine learning to build a model that predicted the point of referral to Emergency Psychiatry (see Methods). We dynamically constructed interaction networks along each patient trajectory separately (i.e., building up the network by adding nodes and edges as each interaction occurred). The state of the network at each point along the patient trajectory was used as input for a Bernoulli naive Bayes classifier [32]. The model was configured to predict the point along a patient trajectory immediately prior to referral. We trained the model on a randomly sampled subset of 80% of the patient trajectories in the data set. To quantify which nodes and edges were most predictive of a referral we used permutation feature importance [33] which was computed based on the balanced accuracy score over the remaining 20% of trajectories. This process was repeated 10000 times for different randomised training and test data sets. The mean balanced accuracy over all training iterations was 82%, 95% CI [59%, 100%].

We found that an interaction between a Registrar (REG) and the patient file, or between a Consultant (CON) and the patient were most predictive of referral on average (Figure 4A and B). The next most predictive events were the involvement of a registrar in the patient trajectory, an interaction between a registrar and the ED Information System (EDIS), and any access of the patient file. The predictive power of all network features had high variance (Figure 4B). For example, the patient file often had a negative value for permutation importance implying that the contribution of this feature to the model could be worse than random. High variance in feature importance is likely a result of the high dimensionality of the feature space relative the number of observations, coupled with the often complex of nature patient trajectories through the ED. However, the model suggests that the involvement of senior doctors in a patient’s trajectory (i.e., a Consultant or Registrar) is more likely to precipitate referral to ED Psychiatry than the involvement of junior doctors (i.e., a Resident Medical Officer (RMO) or Intern (INT)).

**Figure 4:**
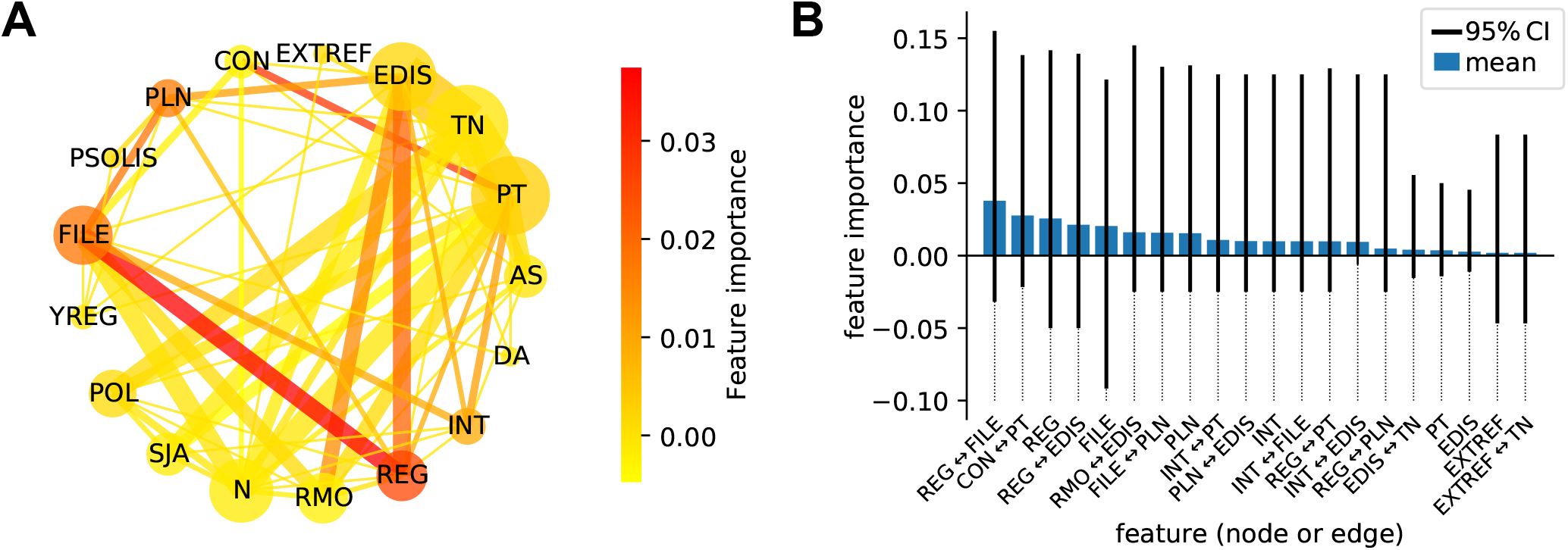
Machine learning on network features reveals agents and interactions that predict the referral from the Emergency Medical team to Emergency Psychiatry: (A) A circular visualisation of the average interaction network for patient trajectories where there is a clinical referral to the Emergency Psychiatry team up to the point of the referral (n=20 patients, n=87 interactions). Node sizes and edge weights reflect the respective relative number of instances for which agents and interactions were observed in this subset of trajectories, where larger size or heavier weight indicates more instances of observation. The color of a node or edge shows its importance for predicting the clinical referral in a subsequent interaction, as computed by permutation feature importance using a Bernoulli naive Bayes classifier trained on the set of nodes and edges at each step of a patient trajectory (see Methods). (B) Mean feature importance and 95% CI for the top 20 nodes or edges over 10000 randomly re-sampled 80:20 train/test splits of the data.

## DISCUSSION AND CONCLUSIONS

This study has introduced a new quantitative framework for investigating the provision of psychiatric care in emergency healthcare settings with a focus on patients with suicide or self-harm risk. By embedding observers in a tertiary hospital emergency department we collected data to construct network models of patient trajectories and clinical interactions respectively. The clinical interaction network had a community structure reflecting the operational division between medical and psychiatric teams. This model indicated that the PLN likely played crucial role in gathering and communicating clinical information between teams, carrying a considerably higher load than other clinicians based on measures of network centrality. Further analysis suggested that this unexpectedly high load may create a risk whereby compromised function of the PLN could lead to reduced information flow between clinicians that negatively impacts patient care. We then used a targeted algorithmic approach to show that this risk might be mitigated by increasing communication between doctors in the Emergency Medical and Emergency Psychiatry teams. Finally we used a machine learning model trained on dynamic network features to identify which clinical interactions were most likely to result in a psychiatric referral.

The unexpectedly high importance of the PLN revealed by our quantitative analysis has implications for operational models that incorporate this or similar clinical roles. PLNs are generally recognised as being beneficial for the provision of mental health care in emergency departments with studies often citing merits such as reduced wait times, positive patient experience, and therapeutic benefits [34, 36, 18, 25, 37, 23, 38, 39, 17, 35, 9]. Notably, qualitative studies have described how PLNs have an important function in communicating information and coordinating patient care included providing assessments and recommendations to doctors, and serving as a link to other hospital services (e.g., alcohol and other drug services) and community mental health services [25, 18]. This is agrees with our observation that the PLN had high centrality in the clinical interaction network. These same studies also reported instances of staff becoming reliant on PLNs and facing considerable impact on workload in their absence [18], and that PLNs can feel unsupported and unsafe due to feeling overloaded with responsibility [25]. These reports are congruent with our finding that high load on the PLN within the interaction network may pose a risk to the function of the system and patient outcomes. This shows that our novel approach appears to capture useful and interpretable information about the implementation of emergency psychiatric care. The advantage of our framework is that it allows for quantification and statistical comparison of different operational models and policies. However, the aforementioned qualitative studies and the present study were undertaken in different hospitals with different operation models. Therefore, future research should seek to validate the network model via a mixed-methods study encompassing both staff interviews and network analysis of clinical interactions in a range of different hospitals and healthcare settings.

We used the clinical interaction network to show that potential system vulnerabilities associated with the high centrality of the PLN might be mitigated by strengthening lines of communication between members of the Emergency Medical and Emergency Psychiatry teams. This echos the more general and well-established finding of the importance of multidisciplinary collaboration and integration for the delivery of effective psychiatric care in emergency settings [25, 38, 27, 34, 40]. Furthermore, a recent study has also identified discrepancies between actual patterns of communication between clinical staff in practice compared with reporting structures as intended in the operational model based on a qualitative study [36]. The quantitative framework we have developed facilitates direct assessment and comparison of actual patterns of communication against policy and organisational expectations in the evaluation of clinical practice. In addition, we have shown how machine learning classifiers can be used in conjunction with the clinical interaction network to understand how patterns of communication impact patient pathways and clinical decision points.

Recent reports highlight the need for new evidence-based measures to evaluate the the implementation of clinical pathways in emergency psychiatry [11, 24, 20, 28], and more data-driven methods for investigating the behavioral aspects of emergency care more broadly, where research is currently limited [41]. The new data-driven framework and quantitative metrics presented here have considerable potential for application and adaption to address a range of challenges in emergency psychiatry. For example, several studies have reported that mental health patients have negative experiences of emergency care due to issues including wait times, lack of appropriate spaces for the provision of care, and negative attitudes of staff during interactions [38, 27, 12, 19]. By incorporating multi-modal data collection of interaction observations, patient interviews, and the appropriate linkage of clinical records, our framework could be extended to evaluate the effectiveness of clinical pathways in terms of patient experience, patient flow, or patient outcomes such as referrals or re-admission rates. In addition, mental health patients who self-present can differ markedly from those brought in by ambulance or police, but details about how their subsequent clinical pathways through emergency care differ are not well understood [42]. Our network-based approach would be ideally suited for mapping, measuring and comparing the nature of patient trajectories for different types of presentations to improve resource allocation, or to develop targeted clinical pathways to enhance treatment outcomes and efficiency for different patient groups.

Future studies utilising our framework may benefit from a larger sample size. This would enable the estimation of transition probabilities along patient pathways, and the frequency of different interactions in the network model to provide a more accurate characterisation of information flow. Deploying additional observers and retrospectively augmenting data using medical records would also reduce the risk of sampling bias during data collection.

In summary, our quantitative framework for mapping patient trajectories in a tertiary hospital emergency department provides a new and complimentary approach for the assessment and improvement of operational models and clinical practice in the provision of emergency mental health care. To conclude, we note that while this study focused specifically on patients with suicide or self-harm risk, our framework could be applied equivalently to investigate other aspects of healthcare service delivery including different medical specialties, other patient groups or demographics, or alternative settings such as community mental health clinics.

## METHODS

### Data collection

Data were collected at a tertiary hospital in the city of Perth, Western Australia. A team of two observers including a Psychiatry Registrar (not on clinical duties) worked simultaneously to record clinical interactions for patients presenting with risk of suicide or self-harm. Observations we taken for a total of 101 hours in the ED over a period of approximately one month, including 40 hours during day shifts, 40 hours during night shifts, and 21 additional hours of observations during day shifts specifically focused around the ED Psychiatry team. Observation hours were distributed between weekdays and weekends, and were generally undertaken in 8 hour shifts. A total n=213 interactions were observed from n=36 patient trajectories in the ED. The Mental Health Observation Area (MHOA) was observed for a total of 17.5 hours over 5 weekdays. A total of n=59 interactions were observed from n=7 patient trajectories in the MHOA.

Observers sought to identify patients for inclusion in the study at the earliest possible opportunity along their trajectory, ideally at the time of presentation. Patients presenting with known or suspected suicidal or self-harm behaviours were approached at an appropriate time and asked if they consented to being observed for the purpose of the study. Observers then followed patients and clinical staff to record all interactions in an event log to the extent that it was practicable (Table 1). This included observations of interactions between the patient, clinical staff, emergency services (i.e., police and paramedics), support and community health services, clinical information systems, and associates of the patient (i.e., relatives or friends). Observations were taken with minimal involvement by the observers who acted as bystanders. However, it was occasionally necessary to briefly interview clinical staff to establish the details of some interactions (i.e., phone calls, access of digital records etc.). No identifying information or clinical information were recorded during data collection.

**Table 1:**
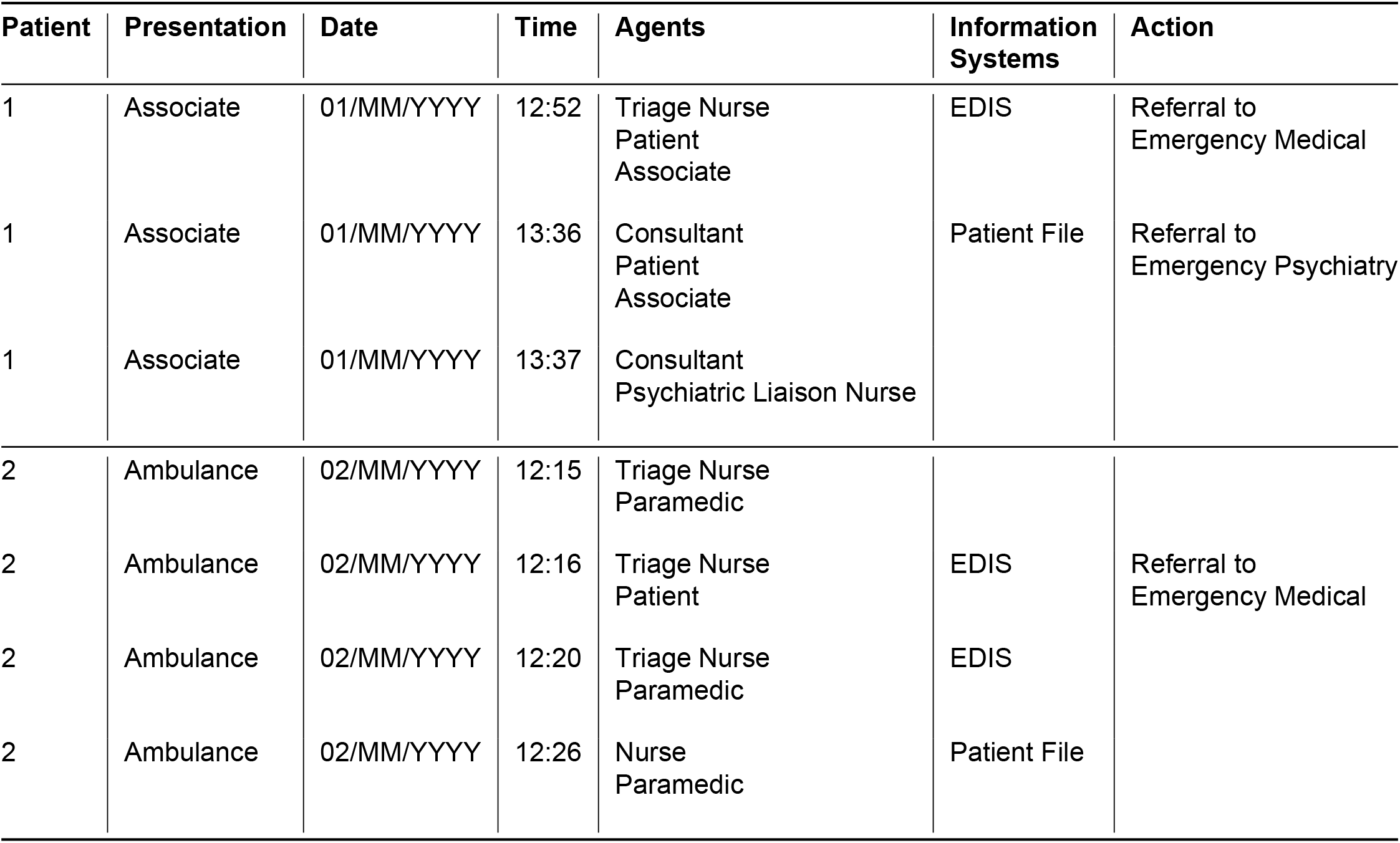
Example event log: Data were collected in the form of an event log similar to this table. Shown here are partial trajectories for two patients. Each row corresponds to an observed interaction, listed in chronological order. Dates have been recorded relative to the date on which data collection commenced.

### Data analysis and software

Data analysis were performed in Python using the packages Numpy [43], Pandas [44], NetworkX [45], Scikit-Learn [46] and Imbalanced-Learn [47]. Figures and data visualisations were prepared using Matplotlib [48] and NetworkX.

### Patient trajectory network

We defined nodes in the network representation of patient trajectories as the clinical team treating the patient, modes of presentation and modes of discharge. An unweighted and directed edge was assigned between a pair nodes if we observed at least one instance of that transition in our data (i.e., a patient being moved from the ED to the Observation Ward). The combined data from all ED and MHOA observations were used to construct the patient trajectory network. When computing the presentation type, trajectory time, interactions, and clinical staff types we excluded patients that were observed only in the MHOA. We excluded this data because the MHOA serves a different function than the other areas of the ED, providing specialised observation of at-risk mental health patients for 24-48 hours. This exclusion was also applied in the subsequent analysis of the clinical interaction network.

### Clinical interaction network

The clinical interaction network is a map of observed interactions between patients, clinical staff, information systems and other agents involved in patient trajectories observed within the ED (see Figure 1D for a complete list of agents and acronyms). The network comprises a set of nodes 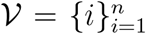, where each node *i* corresponds to one of the |*V*| = *n* total possible agents or information systems. The network is represented by an *n* by *n* adjacency matrix *A*. Elements of *A* are given by *a*_*i,j*_ ∈ {0, 1} where *a*_*i,j*_ = 1 implies an unweighted bi-directional edge between nodes *i* and *j*. An edge was assigned between nodes *i* and *j* if and only if an interaction between the corresponding pair of agents or information systems occurred at least once in the combined data from all patients. The operational policy of a hospital imparts intrinsic structure in the network that is not directly reflected in the event log data. For example, a recorded interaction may involve the patient, a nurse and the patient file which the nurse is either reading or appending information to. However, a patient’s file is never accessed by the patient. Therefore, edges between nodes corresponding to the patient and the patient file are considered forbidden and are excluded from the clinical interaction network by definition. In this study, forbidden edges included those (a) between clinical information systems, (b) between clinical information systems and agents that were not clinical staff at the hospital, (c) between the Psychiatric Services Online Information System (PSOLIS) or ED Psychiatry Handover Document (EDYHO), which are clinical information systems specific to psychiatry, and any agents that were not part of the Emergency Psychiatry team. We rendered the visualisation of the interaction network using the *spring_layout* function for Networkx [45] which produces a force-directed graph layout.

To detect community structure in the network we applied the greedy modularity maximisation algorithm from [29]. Node centrality measures were computed based on the definitions given in [30], as briefly summarised here. The degree centrality of a node is the number of edges connected to that node. The degree centrality of node *i* was computed as:

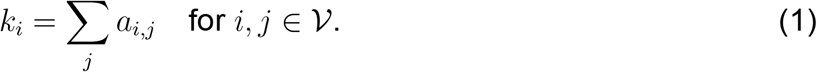

Closeness centrality is the inverse of the average distance from a given node to all other nodes in the network. The closeness centrality of node *i* was computed as:

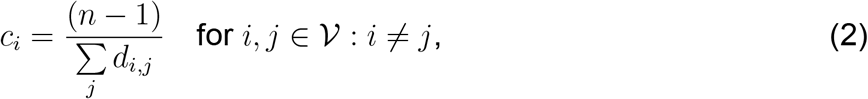

where *d*_*i,j*_ is the length of the shortest path on the network between nodes *i* and *j*. The betweenness centrality of a node measures how often that node forms part of a path between other pairs of nodes. The clinical interaction network models the flow of information between agents. In this context, high betweenness would suggest that a node is important for passing information between other agents or different communities in the network. If a node with high betweenness is compromised this is likely to adversely impact the flow information through the network more than for a node with low betweenness. The betweenness centrality of node *i* was computed as:

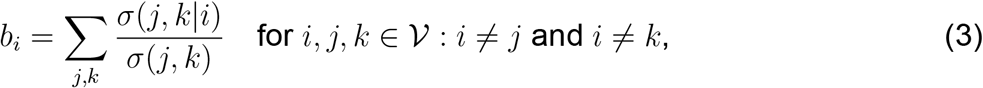

where *σ*(*j, k*|*i*) equals the number of shortest paths between nodes *j* and *k* which pass through node *i*, and *σ*(*j, k*) equals the total number of shortest paths between nodes *j* and *k*. This particular variant of betweenness centrality is described in [49]. We further normalised *b*_*i*_ by the total number of possible paths through node *i* [50].

### Network vulnerability analysis

We applied a random shuffling algorithm to assess the degree to which the structure in our observed network is due to inherent structure rather than randomness. The principle is that we generate an ensemble of networks which appear similar to our clinical interaction network (they have the same number of nodes, node degrees, etc.) but are otherwise random. We then seek to answer the question of whether the observed clinical interaction network is different from random — and if so, how?

Random shuffling of the clinical interaction network was performed using a connected double-edge swap algorithm [31] to preserve local and global degree structure. The algorithm begins by randomly selecting two pairs of nodes (*i, j*) and (*v, u*) such that the nodes within each pair are connected (i.e., *a*_(*i,j*)_ = *a*_(*v,u*)_ = 1). The edges are then swapped so that the network has two new connected node pairs (*i, v*) and (*j, u*). This swap is only performed if: (a) the edges for new nodes pairs (*i, v*) and (*j, u*) did not already exist in the network, (b) the network remains connected after the swap. If these conditions are not met, the edges for these two node pairs are left unchanged and the algorithm proceeds to attempt a swap with a different randomly selected set of node pairs. We impose a further condition that edges can only be swapped if the resulting network remains free of forbidden edges as defined for the clinical interaction network. We generated 1000 shuffled networks from independent sequences of random edge swaps to assess the likelihood of the observed network configuration. For each shuffle we attempted 20000 connected double-edge swaps, of which approximately 1900 swaps were successful on average.

We assessed network vulnerability based on changes in closeness centrality and global efficiency [51] when a potentially vulnerable node was removed. Global efficiency measures how efficiently information propagates on a network. Assuming that efficiency of information flow between a pair of nodes *i* and *j* is inversely proportional to the shortest path between them *d*_*i,j*_, the global efficiency is the average over all node pairs, computed as:

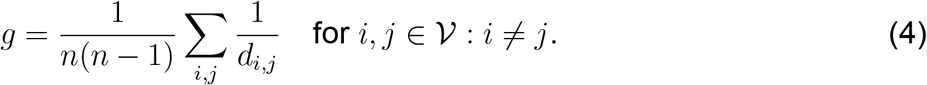

To investigate strategies for mitigating against the adverse effects of a compromised node, we developed a simple greedy algorithm for the addition of edges. The algorithm begins by removing the compromised node from the network. From a set of candidate edges we then added the edge which maximised the increase in global efficiency. This process was repeated until all edges from the candidate set were added to the network. If a tie was encountered with respect to the increase in global efficiency, then the greedy algorithm is no longer guaranteed to find an optimal sequence for the addition of edges. Therefore, once a tie occurred we tested all possible permutations for the sequence of the remaining edges that had not yet been added. This allowed us to enumerate the complete set of optimal solutions. For our data and the specific set of candidate edges investigated in this study there was a subset of edges that were tied with respect to their contribution to global efficiency, regardless of the order in which they were added to the network.

### Machine learning for predicting clinical referrals

To assess which agents and interactions were likely to precipitate a referral to the Emergency Psychiatry team, we trained a binary classifier to predict the referral point based on the evolving state of clinical interaction networks along individual patient trajectories. We used machine learning to achieve this as a way of extracting structural information from the underlying data, independent of our own application driven bias. The machine learning algorithm is agnostic to our knowledge of the system and simply seeks to extract significant structural patterns from the data. We used data from trajectories for n=20 patients who were referred from the Emergency Medical team to the Emergency Psychiatry team during the period of observation. A dynamic clinical interaction network *A*_*p,t*_ was iteratively constructed along the trajectory for each patient *p* and observation number *t* starting with an empty network *A*_*p*_(*t* = 0). For each interaction along the trajectory we added nodes and edges for the corresponding agents if they did not already exist in the network. This process continued up to and including the interaction which precipitated referral. The final state of the *A*_*p,t*_ along the trajectory was assigned a positive class label *y*_*p,t*_ = 1, delineating the the referral point. All other states of *A*_*p,t*_ were assigned the class label *y*_*p,t*_ = 0. Each state of *A*_*p,t*_ was mapped to a 1-dimensional binary feature vector:

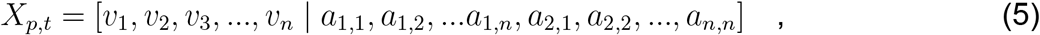

where *v*_*i*_ is a Boolean variable which is true when node *i* exists and *a*_*i,j*_ is the Boolean variable representing the existence of an edge between nodes *i* and *j*. The indices *i* and *j* correspond to those for the complete interaction network *A*.

The features *X*_*p,t*_ and labels *y*_*p,t*_ were then used to train a Bernoulli naive Bayes classifier [32] to predict the referral point based on the dynamic network state. We hypothesised that some features of *X*_*p,t*_ would be more predictive than others, and that it may be useful to identify the agents or interactions corresponding to these features. To investigate this we used permutation feature importance [33] which quantifies the contribution of each feature in the model by measuring the change in a scoring metric when the data for that feature are randomly permuted. We used balanced accuracy [46] as the scoring metric because class labels are highly imbalanced - positive class labels (referral) typically only occur once in a patient trajectory through the ED and only account for 23% of the data. The balanced accuracy is given by:

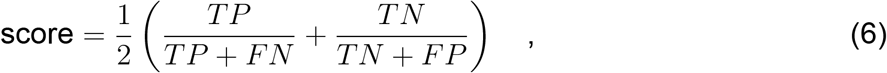

where TP, FP, TN and FN are the number of true positives, false positives, true negatives and false negatives in the test data respectively. We estimated the permutation feature importance for 10000 randomly re-sampled 80:20 train/test splits of the data. Data were grouped such that observations from any given patient trajectory could not be split between the training and test sets. To avoid bias in the model due to highly imbalanced class labels, we used random over-sampling of the minority class label to balance the data in each training split [47].

## Data Availability

Data and code for this study are available online at https://github.com/mhmcc/ED-interaction-mapping

## DECLARATIONS

### Ethics approval

This study was approved as a Quality Activity by WA Health North Metropolitan Health Service and received Ethics Approval from the Chair of the North Metropolitan Health Service Research Ethics Committee. Consent for participation in this study was obtained orally and the data were analyzed anonymously.

### Author contributions

Conceptualization: SH, MS; Methodology: MHM, BJ; Data collection: BJ, MHM; Data analysis & code: MHM; Investigation: MHM, BJ, MS, SH; Writing — original draft: MHM; Writing — review and editing: MHM, MS, BJ, SH; Supervision: MS, SH.

## Acknowledgments

We thank the clinicians and staff at the Sir Charles Gairdner Hospital Emergency Department (North Metropolitan Health Service, Department of Health, Western Australia) several of whom provided advisory and logistical support for this research.

